# Reversible and Noninvasive Modulation of a Historical Surgical Target for Depression with Low Intensity Focused Ultrasound

**DOI:** 10.1101/2024.09.30.24314619

**Authors:** Aki Tsuchiyagaito, Rayus Kuplicki, Masaya Misaki, Landon S. Edwards, Joan A. Camprodon, Kate D. Fitzgerald, Sahib S. Khalsa, Noah S. Philip, Martin P. Paulus, Salvador M. Guinjoan

## Abstract

Major depressive disorder has a point prevalence of 5% of the world population and is the main cause of disability, with up to a third of patients not responding to first-line treatments. Surgical neuromodulation is offered to only an anecdotal proportion of these patients, because while these methods are curative in some individuals, the proportion of responders rarely exceeds 50%. Recent efforts to establish reliable brain circuit-symptom relationships and thus predict response have involved mapping with multiple intracranial electrodes, but the impracticality of this approach currently prevents its application at scale. In the present study (ClinicalTrials.gov identifier NCT05697172; FDA Q220192) we begin to address this gap by leveraging low-intensity focused ultrasound (LIFU), a novel noninvasive technique, to modulate the anterior limb of the internal capsule, which is an established surgical deep white matter target for depression. We based our study on burgeoning in vitro evidence that LIFU attenuates axonal conduction by operating mechanosensitive channels in nodes of Ranvier. Compared with sham stimulus, active LIFU produced a functional disconnection of gray matter hubs reached by the sonicated axonal tracts, an increase in positive emotion, and top-down effects on the cardiovascular autonomic balance. Our results using LIFU of deep-brain white matter tracts in humans open three potential avenues to understand the mechanisms and improve the outcome of depression, namely attaining a personalized definition of brain circuit-symptom relationships, serving as a noninvasive probe for neuromodulation before irreversible procedures in a “try before you buy” approach, and ultimately emerging as a therapeutic intervention itself.

## Main

Major Depression is the main global cause of disability in the employed population^1,2^, and a substantial proportion of patients do not respond to first-line treatments and enter a chronic and incapacitating course, often complicated by suicidal ideation, attempts, and death^3^. These patients are considered to suffer from treatment-resistant depression (TRD)^4,5^. The most severely ill and refractory TRD patients are sometimes offered surgical treatment, via targeted ablation or electrode implantation for focal brain stimulation. These invasive, costly, potentially risky, and largely irreversible procedures target a variety of structures with the hope of interrupting putative dysfunctional circuits that underlie manifestations of the disorder. However, given the point prevalence of depression (5% of the global adult population, or approximately 280 million people, but reaching 6% in women and 5.7% in older adults^6^), individuals receiving these procedures represent just an anecdotal proportion of patients who could potentially benefit from long-term surgical neuromodulation. The impracticalities associated with neurosurgery are the main limitations of these invasive methods. However, there is also modest ability to predict who has chances to respond to invasive methods, given that even in open trials efficacy reaches, with few exceptions, approximately 50%^7^.

Different regions of the anterior limb of the internal capsule (ALIC) have been the most common surgical targets to manage intractable TRD for over half a century^7,8^. This involves the disconnection (by ablation or by localized stimulation via implanted electrodes) of cerebral limbic circuits putatively involved in depression symptoms by targeting large bundles of myelinated axons connecting thalamus, basal ganglia, and brain stem nuclei, with the prefrontal and cingulate cortices. The application of these techniques is empirical, targeting anatomical regions defined at the group level (rather than tailored to each patient’s unique anatomy and symptom profile). This lack of personalization likely contributes to the aforementioned suboptimal outcomes. Precision surgical targeting may be a solution to this challenge, if it could account for the significant interindividual variability in 1) the anatomy of the ALIC and 2) the clinical presentation of TRD. In fact, recent efforts have begun to address this problem by the implantation of electrodes in different brain regions, followed by systematic, individualized probing of circuit-symptom relationships^9^. Nevertheless, at least at this time the invasiveness and cost of this approach limits its application at scale.

In the present double-blinded, randomized, cross-over clinical trial, we begin addressing this problem by leveraging the opportunity offered by low-intensity focused ultrasound (LIFU) to modulate deep white matter tracts reversibly and noninvasively in patients with major depression. We based the development of this study on recent in vitro evidence showing that LIFU disrupts the process of saltatory conduction in myelinated axons^10–13^. This effect is based on the observation that LIFU operates mechanosensitive potassium channels located in the nodes of Ranvier, thus inducing an outflow of cations that results in axon hyperpolarization^14,15^. We hypothesized that applying LIFU to white matter tracts would produce a functional disconnection of the gray matter areas associated to these tracts in a living organism^16^ and reasoned that this disconnection would serve as an in vivo manifestation of the cellular effects observed in previous, in vitro studies. Moreover, we expected that this treatment would improve symptoms of negative emotion in depression (particularly repetitive negative thinking, for the chosen target is also effective in refractory obsessive-compulsive disorder), and normalize the peripheral autonomic responses associated with depression and negative emotion^5–7^. We observed consistent functional disconnection of gray matter hubs linked by the sonicated white matter tracts in the ALIC, along with top-down emotional and visceral changes resulting from functional disruption of such cortico-subcortical limbic circuits. This approach thus opens the possibilities of 1) testing mechanistic links between large-scale brain circuits and depression symptoms (i.e., response biomarkers), 2) probing the function of brain circuits targeted by surgical procedures prior to the definitive intervention^17^ (i.e., predictive biomarkers), and 3) exploring the therapeutic use of LIFU delivered to deep white matter tracts, with intensities that are presumed physically innocuous to the human cerebral tissue (i.e., treatment development)^18^.

## Results

### Noninvasive functional engagement of ALIC network hubs

This study (ClinicalTrials.gov identifier NCT05697172) was determined to be non-significant risk by the Food and Drug Administration (Q220192) and approved by Western IRB. The sample consisted of 21 consecutively recruited participants who had major depressive disorder (MDD) and were stratified according to the intensity of depressive rumination (Figure 1, CONSORT Diagram in Extended Data Figure 1, and %Extended Data Table 1). Each participant was screened, provided consent, and underwent basic clinical and image studies (Table 1). Figure 1a. shows that each participant then underwent an initial experimental intervention visit, a safety visit, a second intervention visit, and a final safety visit, each separated by a week. The two experimental visits consisted of the application of both active and sham LIFU stimuli to all participants, in a randomized order. A CTX-500 SonicConcepts NeuroFUS Pro® device was used to deliver a single 80-s pulsed ultrasound stimulus employing a theta-burst pattern, as previously described^19^. The sham stimulus was identical to the active stimulus, but a 3.175-mm sound-absorbing membrane (Sorbothane®) was interposed between the transducer and the participant’s scalp (Figure 1a). Both study participants and raters were blind to the stimulus condition. The blinding of participants was successful, an important issue given that all emotional and clinical measurements were self-administered (correct detection rate for active LIFU = 50% and for sham LIFU = 67% vs. 50% expected by chance; Χ^2^[1] = 0.59, p = 0.44). Measurements before and after each intervention, including fMRI, are summarized in Figure 1b. The transducer was positioned with the help of a Brainsight® neuronavigator, and the target was defined as the center of a 5-mm radius sphere encompassing the highest number of streamlines connecting the thalamus with orbitofrontal or anterior cingulate cortices in the right hemisphere^20–22^ in each participant, as imaged with probabilistic tractography using MRtrix3^23^. Figure 1c shows the probabilistic tractography image of one exemplar participant. Two separate investigators (AT and SMG) blind to each other’s selection proposed a target for each participant according to this criterion, based upon visual inspection of each participant’s probabilistic tractography image. The two target coordinates thus selected were within 2 mm in 90% of cases, and within 5 mm in all cases. A final consensus on the target coordinates was then agreed upon, before delivering active and sham sonication to each participant. Figure 1d shows the right ALIC target thus defined in all participants, in the common tridimensional anatomical space, reflecting significant interindividual anatomical variability of the white matter tracts in this structure, as reported previously^24^.

**Figure 1.**
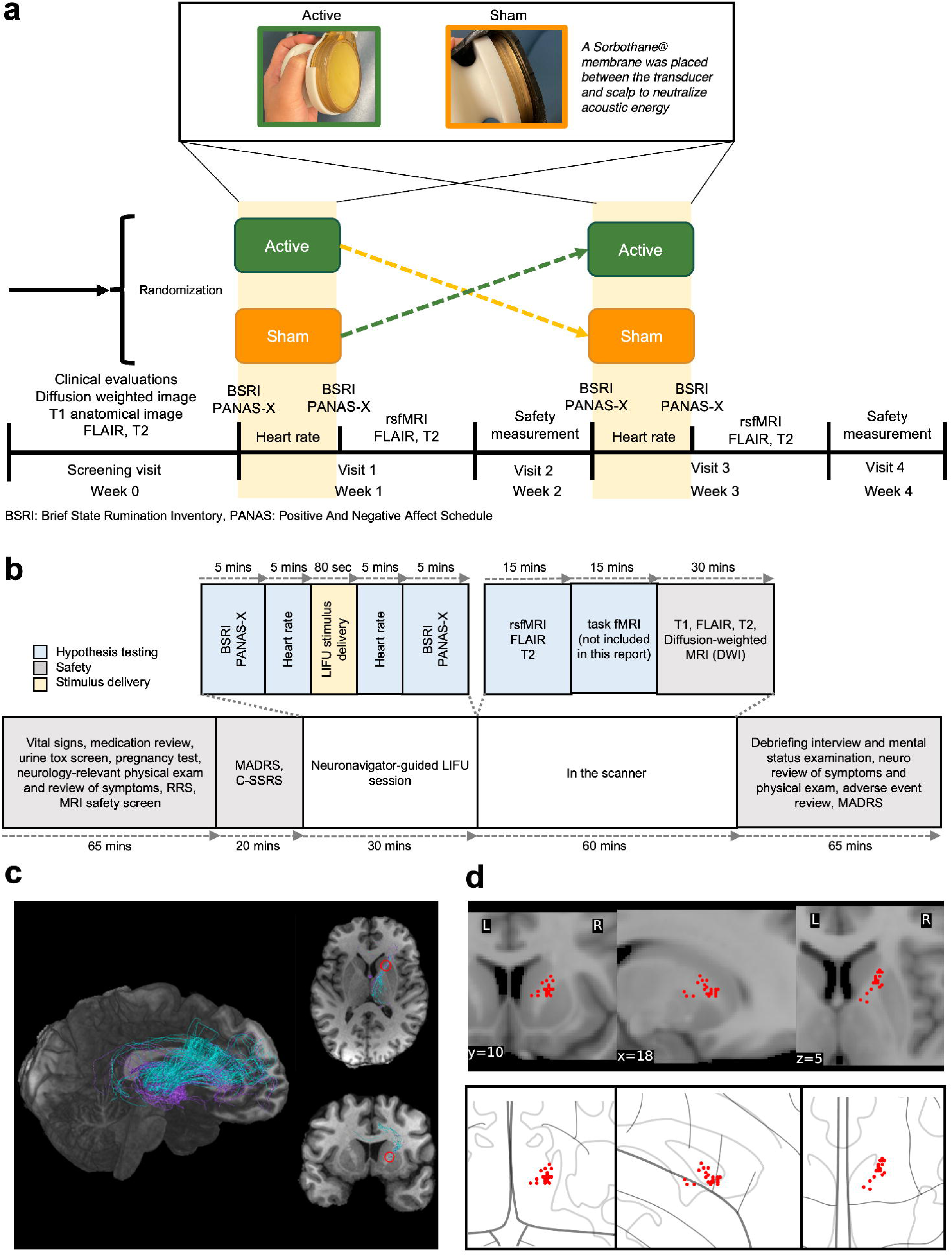
Study design. After a series of screening measures, patients were randomized to receive both active and sham low-intensity focused ultrasound (LIFU) stimuli (a). The sham condition was identical to the active condition but in this case a Sorbothane® membrane was interposed between the transducer and the participant’s scalp. In addition to hypothesis-testing variables, we also measured safety variables including suicidal ideation and behavior, and neurological symptoms and signs, after both interventions, between interventions, and at the end of the study (a). Each interventional visit lasted for approximately 240 min, with safety and hypothesis-testing measurements before and after the 80-second LIFU stimulus (10% duty cycle during a theta-burst pattern, of 20 ms ultrasound every 200 ms; 9.96 W/cm^2^ free-field I_SPPA_), either active or sham (b). The target was defined in each participant, by obtaining a probabilistic tractography of streamlines generated between the thalamus and the orbitofrontal and anterior cingulate cortices and choosing the area with the highest density of these two types of streamlines (c). The dispersion of individual targets in the common MNI space (d) reflects well-described anatomical interindividual variation in the anterior limb of the internal capsule.

**Table 1.**
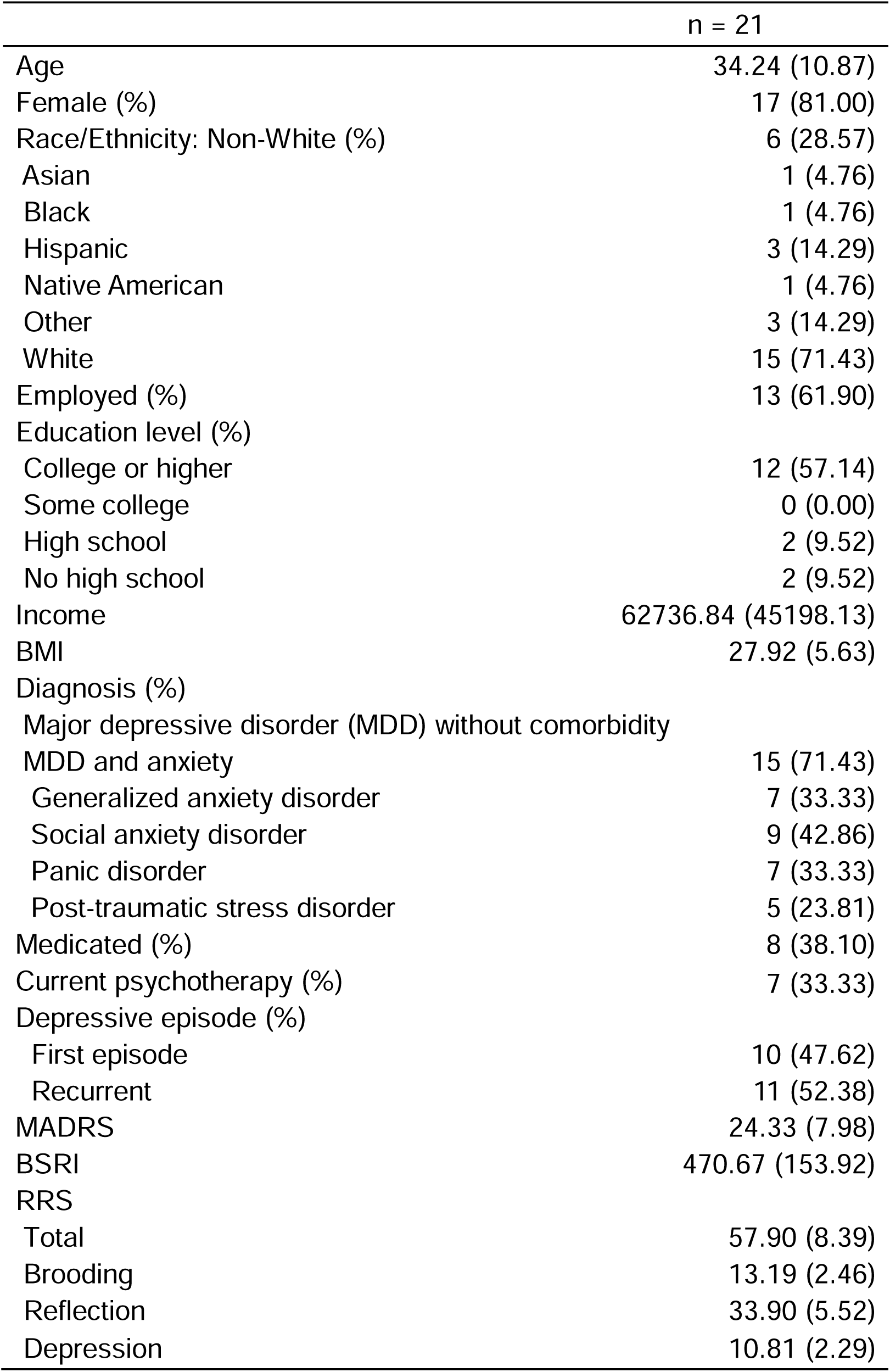
Demographic data.

|  | n = 21 |
| --- | --- |
| Age | 34.24 (10.87) |
| Female (%) | 17 (81.00) |
| Race/Ethnicity: Non-White (%) | 6 (28.57) |
| Asian | 1 (4.76) |
| Black | 1 (4.76) |
| Hispanic | 3 (14.29) |
| Native American | 1 (4.76) |
| Other | 3 (14.29) |
| White | 15 (71.43) |
| Employed (%) | 13 (61.90) |
| Education level (%) |  |
| College or higher | 12 (57.14) |
| Some college | 0 (0.00) |
| High school | 2 (9.52) |
| No high school | 2 (9.52) |
| Income | 62736.84 (45198.13) |
| BMI | 27.92 (5.63) |
| Diagnosis (%) |  |
| Major depressive disorder (MDD) without comorbidity |  |
| MDD and anxiety | 15 (71.43) |
| Generalized anxiety disorder | 7 (33.33) |
| Social anxiety disorder | 9 (42.86) |
| Panic disorder | 7 (33.33) |
| Post-traumatic stress disorder | 5 (23.81) |
| Medicated (%) | 8 (38.10) |
| Current psychotherapy (%) | 7 (33.33) |
| Depressive episode (%) |  |
| First episode | 10 (47.62) |
| Recurrent | 11 (52.38) |
| MADRS | 24.33 (7.98) |
| BSRI | 470.67 (153.92) |
| RRS |  |
| Total | 57.90 (8.39) |
| Brooding | 13.19 (2.46) |
| Reflection | 33.90 (5.52) |
| Depression | 10.81 (2.29) |

Active LIFU resulted in a decrease in functional connectivity between the right thalamus and several cortical destinations of the white matter tracts imaged with probabilistic tractography prior to stimulation, including ipsilateral orbitofrontal cortex and gyrus cinguli, along with homologous contralateral areas, frontal pole, and ventromedial prefrontal cortex (t [19] = −6.89, p < 0.001, d = −1.55) (Table 2 and Figure 2a). The bilateral posterior cingulate cortices were the regions showing maximum functional disconnection after active LIFU of ALIC white matter tracts, compared to sham LIFU (t [19] = −7.26, p < 0.001, d = −1.25) (Table 2 and Figure 2a).

**Table 2.** Significant clusters of the seed-to-whole brain analysis comparing active LIFU vs. sham (right thalamus as a seed).

| **Cluster regions** | **MNI coordinates** | **t-value Active vs. Sham** | **Cluster-mass size** | **Cluster-mass p-FDR** |
| --- | --- | --- | --- | --- |
| vmPFC, Orbitofrontal Cortex, Frontal Pole | -12 +64 +0 | -6.26 | 19921.63 | 0.016 |
| PCC, Precuneus | -4 -40 +30 | -6.71 | 12481.35 | 0.021 |

p-FDR, p-false discovery rate corrected

**Figure 2.**
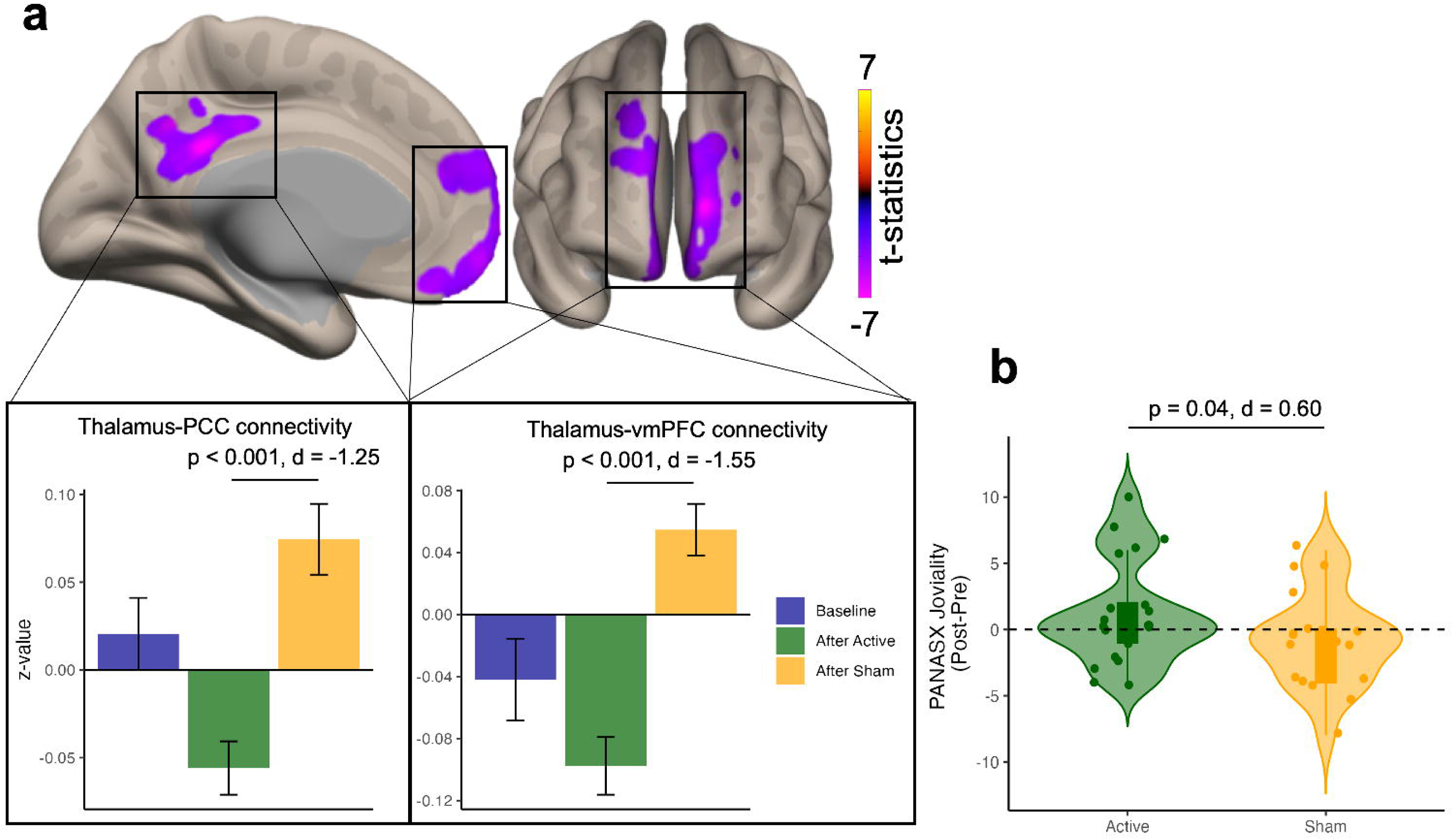
Brain functional connectivity and behavioral results. Active sonication resulted in a reversal of sham sonication-related increased thalamo-prefrontal connectivity. Grey matter destinations of sonicated white matter tracts displaying maximum effect included ventromedial-, medial orbitofrontal- and cingulate cortices (a). A single sonication resulted in an increase in the PANAS-X Joviality subscale, compared to sham sonication (b).

### Emotion and symptom effects of LIFU disconnection of gray matter hubs

Patients were administered the Brief State Rumination Inventory (BSRI^25^) and Extended Positive and Negative Affective Schedule (PANAS-X^26^) scales about 10 minutes before transducer positioning and stimulus delivery, and these measurements were repeated 5-7 min after the stimulus (Figure 1b). Montgomery Asberg Depression Rating Scale (MADRS^27^) depression score was also administered at the end of each experimental session, 2 hours after active or sham LIFU (Figure 1b). There were no measurable changes in ruminative thinking 5-7 min after sonication (Supplementary Figure S1 and %Extended Data Table 2), and no changes in depression severity 2 hours after sonication (Supplementary Figure S1). Most negative affect subcomponents decreased, and positive affect subcomponents increased nonsignificantly after active LIFU compared with sham LIFU (Supplementary Figure S1). Increase in the Joviality subscale of the PANAS-X was statistically significant after active LIFU, compared with sham LIFU (Figure 2b). Greater increase in resting-state thalamo-posterior cingulate cortex (PCC) connectivity after sham conditions was associated increased Attentiveness (r = 0.59, p = 0.006) and Serenity subscales of the PANAS-X (r = 0.509, p = 0.002) (Extended Data Table 3). After active LIFU, greater increase in resting-state thalamo-ventromedial prefrontal cortex (vmPFC) connectivity was associated with a self-reported score of PANAS-X Serenity subscale (r = 0.55, p = 0.013) (Extended Data Table 3).

### Top-down visceral function changes associated with LIFU disconnection of gray matter hubs

A continuous electrocardiogram was obtained beginning 5 min before sonication, throughout the 80-s sonication epoch, and ending 5 min after sonication in all participants (Figure 1b). Heart rate variability (HRV) analysis^28–30^ was applied to the tachogram resulting from consecutive, sinus R-R intervals as described elsewhere^28–30^. We measured HRV reflecting the respiratory sinus arrhythmia (high-frequency, 0.15 to 0.4 Hz, depending solely on vagal influences onto the sinus node) and HRV in phase with intrinsic Mayer waves of the blood pressure (low-frequency, 0.05 to 0.15 Hz, reflecting both vagal and sympathetic influences onto the heart via the baroreflex). The quotient between both types of HRV was then obtained (L/H, reflecting the sympatho-vagal balance onto the sinus node in each individual)^28–30^. Figure 3, panels a and b illustrate this analysis in an exemplar participant (Figure 3a: active LIFU; Figure 3b: sham LIFU). There was an increase in sympathetic relative to parasympathetic sinus node traffic during the sham LIFU session^31^, which was blunted in the active LIFU session (interaction of time and condition: F[2, 95] = 3.77, p = 0.027) (Figure 3c). Regarding specific subcircuits possibly related to this cardiac autonomic activity profile, we observed that greater connectivity between right thalamus and bilateral vmPFC was specifically associated with increased sympathovagal balance in the sham condition (r = 0.46, p = 0.039), and this relationship was lost after active LIFU stimulation (r = −0.20, p = 0.405), possibly at the expense of the decrease in functional connectivity between these structures (Extended Data Table 4 and Extended Data Figure 2a). On the other hand, disrupted connectivity between right thalamus and bilateral PCC after active LIFU was associated with recuperation of cardiac autonomic control as indicated by greater overall resting heart rate variability (SDNN; r = −0.46, p = 0.04) (Extended Data Table 4 and Extended Data Figure 2b).

**Figure 3.**
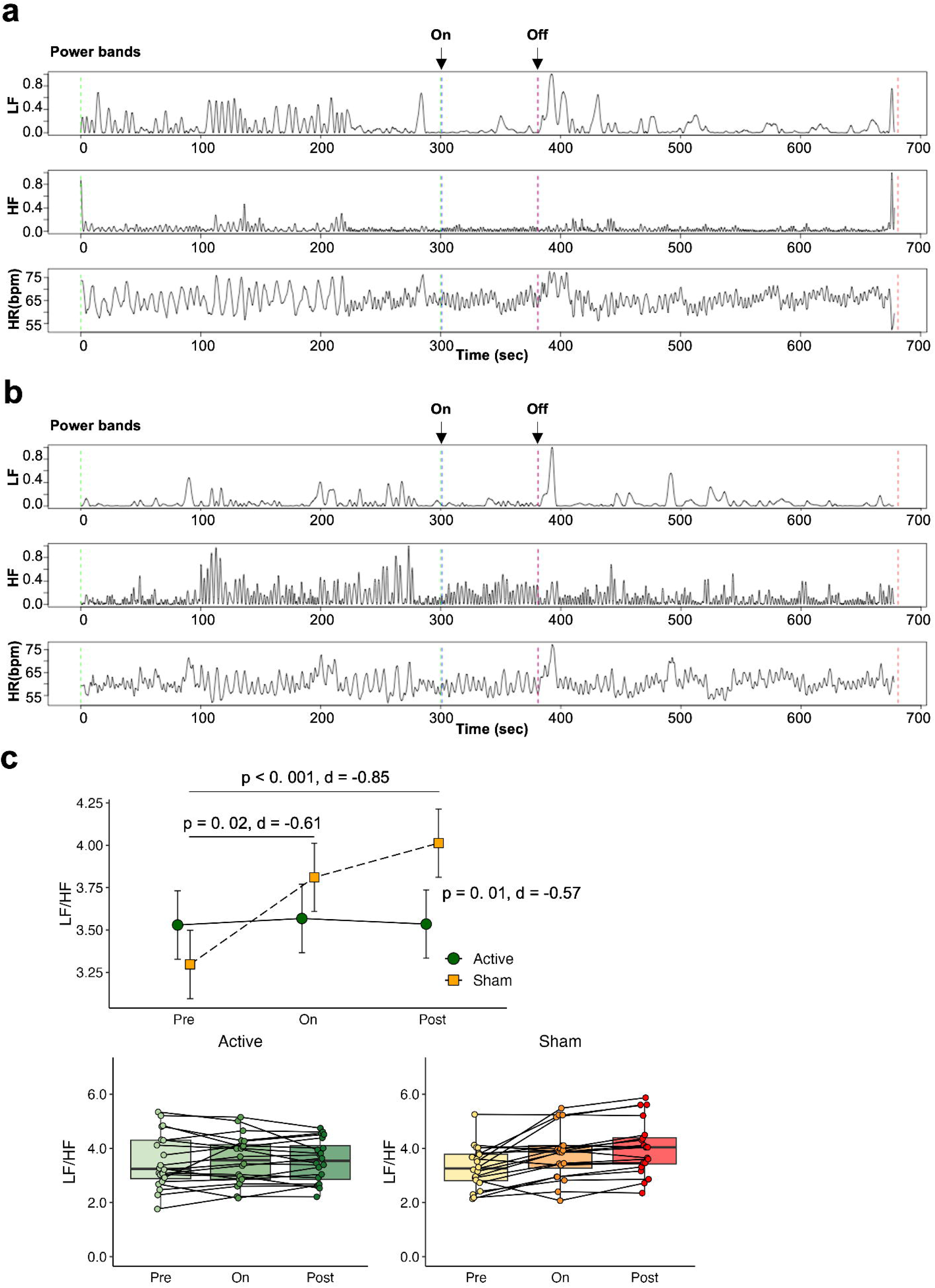
Heart rate variability (HRV) analysis results. HRV was measured during a continuous electrocardiogram recorded 5 minutes before, during, and 5 minutes after sonication. HRV analysis, reflecting respiratory sinus arrhythmia (high-frequency) and intrinsic Mayer waves of blood pressure (low-frequency), was used to determine the sympatho-vagal balance (LF/HF ratio). Panels (a) and (b) show HRV examples for active and sham interventions, respectively. A greater sympathetic response was observed after sham LIFU, which was diminished after active LIFU (c).

### Safety of LIFU of ALIC white matter tracts

All participants underwent a physical exam and DWI and structural T2 and FLAIR sequences at the end of both experimental visits, and in the final visit to detect acute and subacute changes potentially associated with the intervention, in comparison with the same studies obtained at baseline, during the screening visit (Figure 1a,b). A physical and mental status exam was also performed one week after the first experimental visit. Participants did not report adverse effects when specifically questioned. We observed no medical or focal neurological symptoms. Visual inspection of DWI, T2, and FLAIR images did not reveal changes in any patients at all tested moments (not shown).

## Discussion

Applying LIFU to individually-defined white matter tracts traversing the ALIC (a historical surgical target for the treatment of major depression), we demonstrate that it is possible to employ a novel noninvasive, anatomically precise neuromodulation method to produce a functional disconnection of gray matter hubs that participate in the limbic regulation of emotional status and visceral function, and in the pathophysiology of depression^32,33^. The main, acute emotional effect of this intervention is a significant increase in Joviality, an important positive emotion akin to mirth, which has also been observed when the ALIC is acutely engaged by different neuromodulation techniques in the operating room^34–36^. Last, we also observed that the modulation of specific thalamo-corticolimbic regions was associated with an improvement in the sympathovagal input to the heart, such that this balance increased during the sham session -conceivably reflecting a stress response as the intervention was delivered^31^ -and was blunted during the active LIFU session.

We confirmed our initial hypothesis that LIFU of white matter tracts produces a functional disconnection of grey matter regions structurally connected by them. This prediction is based upon extensive in vitro evidence documenting impairment of axonal function by LIFU of comparable intensity. Specifically, we observed a significant decrease in functional connectivity between right thalamus and reciprocally connected orbitofrontal, ventromedial prefrontal, frontopolar, and cingulate cortices. Thus, these results can be best interpreted in light of burgeoning evidence on the effects of LIFU on the electrophysiology of myelinated axons. In vitro, LIFU produces consistent axonal hyperpolarization by operation of mechanosensitive K+ channels in nodes of Ranvier, including TREK-1 and TRAAK channels^10–15^. We speculate that disruption of functional connectivity of grey matter hubs that show reciprocal connections via the white matter tracts targeted in our study can be considered an in vivo mesoscopic network effect of this well-characterized cellular phenomenon, confirming a specific prediction formulated at the time this molecular mechanism of action of ultrasound was discovered^15^.

LIFU has recently shown promise in its ability to allow the testing of mechanistic brain circuit-behavior relationships in health and disease. To the extent of our knowledge, however, extant communications have reported on effects on gray matter only, including the amygdala, cingulate cortical subregions, and thalamus. Nonetheless, some of the most well-established surgical targets for the management of difficult-to-treat psychiatric disorders are located in the deep white matter of the brain, including the ALIC. Furthermore, a major surgical target that was initially defined at the level of cortical gray matter (i.e., Brodmann Area 25^37^), was later observed to owe its therapeutic effects to the pattern of white matter connections engaged by the procedure^38^, leading to the speculation that efficacious long-term neuromodulation is more likely to have widespread symptom effects when it targets large-scale white matter tracts, rather than localized cortical and subcortical grey matter circuit hubs^39^. In this context, our results demonstrating the ability of LIFU to at least transiently modulate functional dynamics across deep white matter tracts effectively and safely in the human brain, may have the potential to inform individual surgical target-symptom relationships prior to ablation or electrode implantation^17^. This potential for translatability to the clinical setting of the present findings of modification of axonal function in humans, can be inferred from the effect of LIFU of white matter tracts in the ALIC on positive affect in our study. Induction of mirth by surgical techniques engaging the ALIC and the ventral striatum has been well documented and is probably the most common acute emotional effect of this type of surgery, providing empirical support of target engagement in our study. Specifically, inhibition of functional connectivity of the cingulate cortex and thalamus, akin to the main outcome of our study, has been described as a distinct neural signature of mirth-inducing DBS^36,40^. To preserve the blinding conditions, in this study raters were not allowed into the venue where sonication was performed, so instant qualitative, subjective behavioral impressions of participants during actual target engagement were not obtained. Nonetheless, the described PANAS-X changes align well with surgical interventions on the target engaged in this study. Further, although the Joviality subscale has received relatively little attention in the clinical literature, it has been recently shown to be associated with improvement in depression and anxiety among patients with mood disorders who are receiving different types of nonpharmacological therapies^41,42^. Whereas in our study we tested the effects of a single sonication, sustained, broader therapeutic effects would likely require several serial stimuli, as it is common practice with all other noninvasive neuromodulation therapies.

Changes in the function of cortico-subcortical circuits as described herein, involving the vmPFC and anterior and posterior cingulate connections with the thalamus, can be expected to exhibit autonomic changes given their well-documented, top-down visceral regulatory role, as part of the central autonomic network^43–46^. This issue is important in patients with major depression, in whom dysfunction of visceral effector organs is a prominent clinical component -to the extent that more patients with depression are seen by cardiologists, gastroenterologists and general practitioners than by psychiatrists^47^. Commonly measured through standard tests of autonomic function, including heart rate variability analysis, autonomic dysfunction is a well-established phenomenon in depression, especially in cardiovascular territories^48–54^. It manifests in the form of increased sympathetic output, decreased parasympathetic output, or both. This visceral disturbance profile is actually considered a major pathophysiological mechanism for the appearance of heart disease and its complications in patients with depression, as well as a poorer prognosis in patients with cardiovascular diseases who become depressed^55,56^. Our finding that LIFU of the ALIC blunted the increase of sympatho-vagal balance vis-à-vis the testing situation^31^, provides an easily measurable, albeit indirect form of assessing target engagement and possibly, beneficial top-down visceral consequences of the disconnection of circuits responsible for this important bodily component of depression. This observation is also in alignment with results by our group and others, describing acute adjustments in the autonomic nervous system function profile in relation to symptom changes in the context of deep-brain stimulation for depression^56–58^, including those in which the ALIC is surgically targeted^58^. We are mindful these results were driven by effects in the sham group and as such, replication is needed.

There are several limitations that need to be considered. First, patients participating in the study were ambulatory and not selected for treatment-resistance, which is the condition that can be managed with ALIC neuromodulation. Second, we chose an individualized target for each participant, seeking to account for interindividual variability in white matter tract disposition. Whereas this may have resulted in improved homogeneity of functional changes across individuals, most surgical approaches target a structure defined in the common anatomical space (e.g., ventral capsule/ventral striatum), which again makes our findings not immediately generalizable to the usual surgical neuromodulation interventions. Third, whereas functional brain tissue engagement strongly suggests we were able to focus the ultrasound beam on the proposed target, we do not have confirmation of direct tissue effects in the sonication target with the available technology. This is an active area of investigation, and emerging data on MRI sequences like arterial spin labelling^59^ may be helpful to confirm the location of the focused ultrasound beam in the future. Also, for safety reasons we have chosen to use the same free-field energy in all participants, as described before^60^. However, cranium thickness and shape may influence the final energy values of the LIFU stimulus in the deep target tissue, thus introducing significant interindividual variability in the actual exposure to LIFU. Given that the intervention was a single application, whether and how these findings (and safety) replicate with repeated LIFU administrations remain unknown.

These limitations notwithstanding, the present results provide evidence demonstrating that deep white matter tracts of the human brain can be safely and effectively engaged with the application of LIFU. This is a novel cost-effective and noninvasive tool, whose application to a historical surgical target useful in the treatment of depression resulted in consistent functional disconnection of grey matter hubs, acute emotional changes, and effects in peripheral sympatho-vagal output. The potential future applications of this observation are manifold. First, LIFU can be systematically applied to discover causal relationships between anatomically defined large scale circuits and symptoms. The use of LIFU in large groups of individuals (which is not possible or practical with invasive techniques, including the implantation of multiple electrodes to test different circuits in a single patient) may thus result in the confirmation of individual targets for treating depression symptoms. Similar to the stepwise cardiac electrophysiology approach to mapping, reversible probing, and ultimately ablating supraventricular arrhythmias^61^, LIFU could also serve the purpose of probing circuits in individual patients prior to a definitive surgical procedure, hopefully increasing the chances of clinical response^17^. Lastly, it may be possible to envision a future where LIFU itself can be tested as a treatment, especially if applied in several sessions to produce neuroplastic changes^18^, as it has been observed in the case of other non-invasive neuromodulation techniques. In this scenario, the advantage of LIFU is the ability to deliver precise, focal and selective stimulation to deep in limbic circuits involved in the formation of mood disorder symptoms (something not possible in the case of traditional repetitive transcranial magnetic stimulation, rTMS). By describing neuroimaging, behavioral, and physiological indicators of acute LIFU engagement of deep white matter tracts in humans, this study provides the initial foundation upon which to begin addressing those research tasks and clinical applications.

## Data Availability

External researchers can make written requests for data sharing. Requests will be assessed on a case-by-case basis in consultation with the lead and coinvestigators.

## Methods

### Trial design

In this double-blinded, randomized, cross-over clinical trial, we aimed to evaluate the effects of LIFU on the modulation of deep brain circuits in individuals with major depressive disorder. Specifically, this study was designed to target white matter tracts in the ALIC of the right hemisphere^20–22^, a well-established surgical site for TRD. Our primary objective was to determine if noninvasive LIFU could effectively disrupt functional connectivity in ALIC-associated gray matter hubs, while assessing its impact on repetitive negative thinking in patients with depression. To disentangle any potential specific effects on perseverative mentation independent from emotion changes, we also evaluated acute changes in negative and positive emotion with the PANAS-X scale, along with cardiac autonomic changes. Last, general symptoms of depression and safety scores including a suicidal thinking and behavior scale, a neurological physical exam, and structural MRI sequences were obtained at the end of each testing session, as specified below.

The study employed a personalized approach to target selection, beginning with probabilistic tractography to individually define the target white matter tracts in each patient, followed by delivery of both active and sham LIFU stimuli in a randomized sequence. Both participating patients and raters were blind to the type of stimulus. After initial participant screening and clinical evaluation, all patients underwent two separate intervention visits, one week apart, where both active and sham LIFU were delivered in random order. Participants were blinded to the intervention conditions, as were the raters, and all outcome measures were collected under blinded conditions.

### Regulatory information and safety monitoring

The study protocol was approved by the Food and Drug Administration under a Non-Significant Risk (NSR) determination (Q220192) and received ethical approval from the Western Institutional Review Board (WIRB Protocol #2021-011-00) for clinical testing. The trial was registered on ClinicalTrials.gov (NCT05697172). All study participants provided written informed consent in accordance with the principles outlined in the Declaration of Helsinki. Informed consent was obtained by trained research staff, and participants were fully informed about the nature and purpose of the study, including potential risks.

Safety monitoring was conducted at multiple time points, including an initial screening, both intervention visits, a safety visit one week after the initial intervention (active or sham LIFU), and a final follow-up visit one week after the second intervention. Structural imaging, such as T2 and FLAIR sequences, was conducted at the screening and after each intervention to monitor for any acute or subacute changes. Physical exams and mental status evaluations were also performed to assess for neurological symptoms or adverse events, and any unexpected findings were reported to the independent Data and Safety Monitoring Board (DSMB).

The DSMB, composed of one neuromodulation expert, one statistician, and one clinical psychiatrist, oversaw participant safety throughout the study. The DSMB was authorized to pause the trial in response to any unexpected adverse events and had the authority to terminate the study if a second unanticipated event occurred, such as neurological complications or tissue changes related to LIFU application. Adverse events were monitored, and the safety assessments ensured that no participants experienced structural changes or significant health risks during the study.

### Participants

All patients were recruited from participants in the Laureate Institute for Brain Research (LIBR) Centers of Biomedical Research Excellence (CoBRE) study (WIRB Protocol #20182352) between December 2022 and February 2024. Twenty-one outpatients with major depressive disorder (MDD) and varying intensities of repetitive negative thinking (RNT) were enrolled. All participants were selected based on their Ruminative Response Scale (RRS)^62^ scores, which was used to determine their baseline levels of RNT. The sample was evenly distributed between patients with low (RRS<60) and high (RRS≥60) baseline trait RNT (see Extended Data Figure 1), given this is a symptom improved by surgical neuromodulation of the ALIC, and we hypothesized active LIFU to affect RNT. Patients were included if they met the following criteria: a Patient Health Questionnaire-9 (PHQ-9)^63^ score ≥ 10 at enrollment, willingness to comply with all study procedures, and availability for the study duration. They were also required to be aged 18–60, in good general health, and able to provide informed consent. Female participants of reproductive potential underwent a urine pregnancy screening test to ensure eligibility. Participants were excluded if they had a history of schizophrenia spectrum disorders, non-affective psychotic disorders, or bipolar disorders. Additional exclusion criteria included a lifetime history of drug abuse, presence of any MRI contraindications, febrile illness, or treatment with an investigational drug within the past two weeks. Suicidal ideation or behavior, determined by the Columbia-Suicide Severity Rating Scale (C-SSRS)^64^, was also an exclusion factor (Extended Table 1). All patients continued to receive care from a licensed physician or mental health care provider throughout the study, but no changes of psychotropic medications were allowed since 6 weeks prior to enrollment and throughout the study participation.

### LIFU intervention

This pilot study included 21 participants diagnosed with MDD. After informed consent was obtained, participants were randomly assigned to receive either LIFU or sham first in a cross-over design with a blocked randomization with a block size of two (High or Low RNT). Interventions were delivered by two of the investigators (AT and SMG) who did not participate in the evaluation of participants, including hypothesis-testing and clinical safety evaluations. Structural MRI were screened for abnormalities in a blinded manner as well. Both participants and the team conducting efficacy and safety assessments were blinded to the treatment condition, though the operators delivering the intervention were not. The sham condition was implemented by placing sound-absorbing material (Sorbothane®) between the transducer and the participant’s scalp, which was applied to a glabrous skin region in the vicinity of the union between the frontal and temporal bones on the right forehead.

Before randomization, all participants underwent the following baseline neuroimaging assessments: a T1-weighted structural MRI for neuronavigation and accurate positioning of the LIFU device, 6-min resting-state fMRI to establish baseline functional connectivity, which was compared to post-treatment scans to evaluate connectivity changes, and multishell DWI that was used to define structural connections, which guided LIFU focus using neuronavigation software (Figure 1). We report fMRI data on 20 patients (1 excluded due to excessive head motion as explained in Anatomical and functional MRI data preprocessing section), symptom data on all 21 patients, and HRV data on 20 patients (one excluded due to >10 premature beats in each 5-min, pre- and post-intervention periods).

### Neuromodulation procedure

#### Individual definition of the neuromodulation target

As a first step to define the sonication target, probabilistic tractography was used, as previously described^23^, to characterize thalamo-orbitofrontal and thalamo-anterior cingulate streamlines in each individual patient. Tracts connecting the right thalamus with both orbitofrontal or anterior cingulate cortices were defined as previously employed by our group and described in detail elsewhere^23^. Briefly, we used MRTrix3 software implemented in Matlab to select the corresponding grey matter seeds according to the Desikan-Killiany brain atlas^65^, and then to estimate structural streamlines connecting those pairs of structures, employing a probabilistic tractography analysis^23,66–74^. Groups of streamlines were then targeted by active and sham LIFU, as explained above. The stimulation focus was centered in the tridimensional coordinate engaging the largest number of streamlines, such that a 5-mm radius sphere whose center is located at the coordinate, will engage the highest possible number of streamlines for either type of connection upon visual inspection of the tractography image. The definition of the sonication focus by two separate investigators blind to each other’s choice (AT and SMG) resulted in targets that were within 2 mm for 90% of subjects, and within 5 mm 100% of subjects, i.e., within the diameter of the expected neuromodulation focus. The final sonication target was then agreed jointly by the two investigators.

#### LIFU Positioning and Stimulus delivery

LIFU was administered using a CTX-500 NeuroFUS Pro device (SonicConcepts Inc.), targeting the ALIC to modulate thalamo-orbitofrontal and thalamo-anterior cingulate white matter tracts. We used an 80-second train of 20-millisecond bursts of ultrasound (0.5 MHz), repeated every 200 milliseconds (for a total of 400 bursts), which represents a 10% duty cycle, and a total energy of 9.06 W/cm^2^ ISPPA. These proposed stimulation parameters^19^ result in estimated tissue values for ISPPA = 2.26 W/cm^2^ and ISPTA = 0.22 W/cm^2^ in a focus of approximately 1 cm^3^, considering an approximate 75% attenuation due to scalp and bone transmission^75^. When assuming a linear free field, we calculated a pressure of 259 kPa with ISSPA set to 2.26 W/cm^2^. This stimulation intensity is lower than the limits recommended by the FDA for diagnostic ultrasound, namely ISPTA ≤ 0.72 W/cm^2^ and ISPPA ≤ 190 W/cm^2^, respectively^76^. Our study accordingly received a Non-Significant Risk (NSR) determination by the Food and Drug Administration (FDA). The administration of LIFU was near the union of the frontal and temporal bones in all participants, which allowed for the closest accessibility to the white matter targets to sonicate and had the advantage of delivering the stimulus in glabrous skin, thus minimizing the presence of any air bubbles in the ultrasound conducting gel. Since the free-field ISSPA maximum limit of the NeuroFUS® system is fixed at 30 W/cm^2^ (free field pressure at 30 W/cm^2^ = 944 kPa), i.e. higher than the intended stimulus, to minimize the chances of errors during the administration of the stimulus, all subjects underwent LIFU under identical conditions, as previously published^60^. These conditions were fixed with the “Save” option in the operator interphase. In addition, to prevent accidental changes in the parameters of stimulation, the two operators independently verified that the parameters were as defined in the protocol, prior to each stimulus administration. The depth of the ultrasound focus was instead steered to match the depth of the target as indicated by a BrainSight® neuronavigation system, which was also used to position the ultrasound transducer.

#### Safety Assessments

Prior to each session, participants underwent a neurological examination. Imaging scans, including FLAIR, DWI, and T2, were used to confirm the safety of the intervention. Baseline and post-sonication scans were compared to detect any adverse effects, such as structural changes in the brain.

#### Clinical hypothesis-testing assessments

All participants were tested with scales to assess acute state-rumination and emotional changes, which were administered 5-10 min before and 5-8 min after actual sonication. In all cases, the Brief State Rumination Inventory was administered immediately before the Positive and Negative Affect Schedule.

##### Brief State Rumination Inventory (BSRI)^25^

This is a validated scale to assess the momentary status of participants, assessing state rumination with 8 questions beginning with “Right now (…).”

##### Positive and Negative Affective Schedule-Extended (PANAS-X)^26^

This is a well validated and extensively used self-report questionnaire consisting of two scales to measure both positive and negative affect, rated on a 5-point scale.

#### Group-level analysis

Group-level analysis of clinical variables was conducted by calculating the post-pre values for the BSRI and PANAS-X subscales. Wilcoxon signed-rank tests were used to compare the post-pre differences between the active and sham sessions. Additionally, Z-values were extracted from the significant clusters based on the seed-to-whole brain analysis, and correlations between functional connectivity and clinical variable changes (post-pre) were analyzed using Pearson’s correlations (p-values uncorrected for multiple comparisons).

#### Additional clinical variables

##### Columbia-Suicide Severity Rating Scale (C-SSRS)^64^

Suicidal ideation and behavior were monitored pre- and post-intervention for safety purposes. If clinical significance was detected, a psychiatrist conducted a follow-up evaluation and determined whether acute intervention was necessary.

##### Montgomery-Asberg Depression Rating Scale (MADRS)^27^

This standard depression symptom scale was applied in all visits at the end of each visit to observe general depression symptomatology changes in relation to the intervention.

##### Ruminative Responses Scale (RRS)^62^

The RRS is the most commonly used score system to assess the intensity of trait rumination. This was employed upon screening to allocate study participants to groups with high (RRS≥60) or low (RRS<60) baseline repetitive negative thinking.

### Functional imaging hypothesis-testing assessments

#### Neuroimaging data acquisition and preprocessing

All the participants went through an anatomical scan and then two 6-minute resting-state fMRI scans were obtained subsequently, separated by a short break^77^. We report on the initial 6-min epoch. During the resting-state fMRI scan, participants were instructed not to think about anything in particular, and to be relaxed while looking at a cross on the screen. For the anatomical reference, T1-weighted MRI images with a magnetization-prepared rapid gradient-echo (MPRAGE) sequence were acquired with the following imaging parameters: FOV = 256 mm, matrix = 256 × 256, slice thickness = 1.0 mm producing 1mm isotropic voxels, 208 sagittal slices, TR/TE = 6/2.92 ms, SENSE acceleration factor *R* = 2, flip angle = 8°, inversion time TI = 1400/1060 ms, sampling bandwidth = 31.25 kHz, scan time = 6 min 11 s. For the resting-state fMRI, a single-shot gradient-recalled echo-planner imaging (EPI) sequence with sensitivity encoding (SENSE) was used with the following imaging parameters: TR/TE = 2000/27 ms, flip angle = 78°, FOV/slice = 240/2.9 mm, 39 axial slices, SENSE acceleration factor *R* = 2, 96 × 96 matrix reconstructed into 128 × 128 image resulting into 1.875 × 1.875 × 2.9 mm^3^ voxels volume, sampling bandwidth = 250 kHz, scan time = 6 min (180 TRs).

#### Anatomical and functional MRI data preprocessing

Imaging data process were performed with SPM12 (Wellcome Department of Imaging Neuroscience, UCL, London, UK), CONN-toolbox (version 20.b)^78^, implemented in Matlab R2019a (The MathWorks Inc., Natick, MA, USA). The initial three volumes were discarded to avoid magnetic saturation effects. The default preprocessing pipeline implemented in CONN-toolbox was performed for both anatomical and functional imaging. Those preprocessing pipeline included functional realignment and unwarp, slice-timing and motion correction, outlier detection, segmentation of gray matter (GM), white matter (WM) and cerebrospinal fluid (CSF), normalization to Montreal Neurological Institute (MNI) space, and partial smoothing (6 mm full width at the half maximum Gaussian kernel). During the outlier detection step, acquisitions with framewise displacement (FD) above 0.25 mm or global mean intensity above three standard deviations were flagged/scrubbed as potential outliers using the Artefact Detection Tools (ART: www.nitrc.org/projects/artifact_detect). If the number of volumes flagged as outliers divided by the total number of volumes was greater than 25%, that subject was excluded from the analysis. After the preprocessing, denoising of the functional data was performed using a component-based noise correction method, (CompCor) by taking into account five orthogonal time series and their derivatives from WM and CSF^79^, linear detrending to remove the linear signal drift, temporal band-pass filtering (0.008-0.09 Hz) to remove physiological, estimated subject-motion parameters (3 rotation and 3 translation parameters and 6 other parameters representing their first order time derivatives), and scrubbing parameters derived from the ART.

#### Seed-to-whole brain connectivity analysis

Following the preprocessing steps, we generated whole-brain connectivity maps for each participant across all sessions. The Brainnetome thalamus sub-regions parcellation atlas^80^ was utilized to define seed regions, particularly focusing on the regions of the thalamus known to have reciprocal connections to the prefrontal and cingulate cortices. We observed that the pre-motor thalamus as defined in the Brainnetome atlas^80^ showed statistically significant connectivity in relation to the experimental intervention. This large region (Supplemental Figure S2), encompasses diverse anatomical thalamus nuclei including anterior, ventrolateral, ventromedial, ventral posterior-medial, and ventral posterior-lateral nuclei, which are known to have reciprocal connections with widespread areas of the prefrontal and cingulate cortices^86^. A connectivity map for this seed was created by correlating the averaged time series within the seed mask to all other voxels in the brain. These connectivity values were transformed into z-scores using Fisher’s transformation for standardization. To compare connectivity differences between the sessions (active vs. sham LIFU), paired t-tests were performed. Given the relatively small sample size (n = 21), we employed a permutation/randomization-based cluster-level inference method^81^ to improve sensitivity while maintaining error rate control. Initially, voxel-wise statistical maps were thresholded at an uncorrected p < 0.01 to identify clusters of interest. Significant clusters were then refined using a cluster-level threshold corrected by False Discovery Rate (FDR) at p < 0.05. This approach allowed for the identification of meaningful clusters while controlling for false positives.

#### Autonomic Hypothesis-Testing Assessment^28,29,83^

We obtained a continuous DII or aVF electrocardiogram (ECG) signal with a distinct R-wave beginning 5 minutes before, and finishing 5 minutes after the intervention, for a total of approximately 12 minutes including the 80-sec LIFU application epoch (BIOPAC Systems, Inc., USA). ECG was measured using three disposable pre-gelled EL258RT electrodes placed on the left chest area: positive at V5, negative at V2, and neutral between them. These electrodes were connected to the ECG100C amplifier. Signals were recorded at a sampling rate of 1 kHz and processed using the BIOPAC AcqKnowledge software, with additional analysis employing wavelets as previously described^83^, and performed using R package RHRV^84^ with R version 4.2.2.

#### HRV analysis

Data preprocessing was carried out to load and filter the heartbeats, compute instantaneous heart rate (HR), and remove outliers. The heartbeat positions were thus filtered to remove outliers and spurious points that resulted from artifacts or undetected heartbeats, and were individually inspected in all cases. To facilitate spectral analysis, the instantaneous HR series was interpolated to achieve a uniformly spaced dataset^83,84^. Linear interpolation at a frequency of 4 Hz was applied.

#### Time-Domain Analysis

Time-domain analysis was performed to calculate the Standard Deviation of all normal sinus rhythm RR intervals (SDNN)^28^. A window size of 300 seconds and a bin interval of 7.8125 ms were used to generate these parameters.

#### Frequency-Domain Analysis^28,29,83,84,85^

The RHRV^84^ package employs an adaptive threshold filter (default settings) and calculates spectral power via the Maximal Overlap Discrete Wavelet Packet Transform (MODWPT^85^). The wavelet-based spectral method in the RHRV package calculates power within predefined spectral bands, using a specific tolerance to address band boundary issues. Power was calculated for the Low Frequency (LF) (0.05-0.15 Hz) and High Frequency (HF) (0.15-0.40 Hz) bands, in addition to the LF/HF ratio. We used an absolute tolerance of 0.01 Hz, and the least asymmetric Daubechies (d4) for performing the wavelet analysis^84^.

#### Group-level analysis

Linear mixed-effects (LME) modeling was performed, with time (pre, on, post) and session (active vs. sham) as interacting random effects and subject as an intercept. Similar to clinical variables, correlations between functional connectivity and changes in HRV indices (post-pre) were calculated, and we report the uncorrected for multiple comparisons p-values.

## Acknowledgements

This work was supported in part by the Laureate Institute for Brain Research, and the National Institute of General Medical Sciences Center Grant Award Number (P20GM121312, PI: MPP) as was the efforts of the authors: JAC (R21AG078692, R61MH132869, R21AG081925, R21MH131878), KDF (R33MH121641, R61MH129544), SSK (R01MH127225), NSP (I50 RX002864, U01 MH123427). The views expressed in this article are those of the authors and do not necessarily reflect the position or policy of the Department of Veterans Affairs or National Institutes of Health.

---

**Extended Table 1.** Inclusion and exclusion criteria.

|  |
| --- |
| ***Inclusion criteria*** |
| A diagnosis of Major Depressive Disorder |
| A PHQ-9 score $\geq 10$ at enrollment. |
| Provision of signed and dated informed consent form. |
| Subject provides verifiable contact information (name, telephone number(s), email and mailing address) for at least 2 persons who agree to be contacted by study personnel as deemed necessary. |
| Subject is followed by a licensed physician or a licensed mental health care provider (i.e., psychologist, LCSW) throughout study participation outside of LIBR. |
| Stated willingness to comply with all study procedures and availability for the duration of the study. |
| Male or female, aged 18 to 60 years. |
| In good current general health as evidenced by medical history. |
| Ability to comply with the study chronogram. |
| For females of reproductive potential: negative urine pregnancy screening test. |
| ***Exclusion criteria*** |
| Current use (within the last 30 days) of drugs of abuse or moderate / severe alcohol use disorder. |
| Lifetime diagnosis of schizophrenia spectrum disorder, other nonaffective psychotic disorders, or bipolar disorders. |
| Presence of cardiac pacemaker or any other MRI contraindication. |
| Pregnancy or lactation. |
| Febrile illness within the last two weeks. |
| Treatment with an investigational drug or participation in any other interventional research protocol in the last 2 weeks. |
| The participant is unable to understand the goal of the study, instructions, or the risks associated with the study as judged by a clinically trained assessment team member. |
| Clinical and/or imaging evidence of vascular, traumatic, or neurodegenerative disorders of the CNS, or other neurological disorders potentially compromising patient's participation in the study, or study results. This includes, but it is not limited to, any minor or major neurocognitive disorder including those caused by traumatic brain injury, Parkinson's disease, significant small-vessel disease, multiple sclerosis, Huntington's disease, early-onset Alzheimer's disease, chronic infections of the CNS or the meninges, previous chronic use of alcohol or CVA sequelae, or a Montreal Cognitive Assessment (MoCA) score $<25$ due to any cause, to be obtained if cognitive dysfunction is suspected. The PI can decide if a potential participant needs to be excluded due to some other cause of structural or functional compromise of the CNS (e.g., epilepsy). |
| Active suicidal ideation (as measured by Suicide-Risk-Assessment-C-SSRS "Yes" answers to items 3, 4 or 5 of Suicidal Ideation-Past 1 month section, or any "Yes" answer to any of the items of Suicidal Behavior-Past 3 months section), or any suicide attempt in the last year. |

**Extended Data Table 2.** Changes in symptoms and mood and statistical tests.

|  | Active |  |  | Sham |  |  | Wilcoxon signed rank test |  |
| --- | --- | --- | --- | --- | --- | --- | --- | --- |
|  | Pre | Post | Post-Pre | Pre | Post | Post-Pre | p-value | Effect size |
| **BSRI** | 442.00<br>(162.12) | 360.19<br>(199.28) | -81.81<br>(103.66) | 424.62<br>(192.85) | 337.62<br>(189.84) | -87.00<br>(116.14) | 0.578 | -0.14 |
| **PANASX** |  |  |  |  |  |  |  |  |
| **Negative affect** | 20.48 (8.45) | 15.86 (7.07) | -4.62 (4.08) | 18.81 (8.69) | 15.95 (7.76) | -2.86 (4.76) | 0.304 | -0.10 |
| **Positive affect** | 18.81 (6.10) | 19.43 (5.45) | 0.62 (4.28) | 20.86 (6.87) | 20.76 (6.89) | -0.10 (3.06) | 0.518 | 0.26 |
| **Fear** | 11.43 (5.40) | 9.67 (5.38) | -1.76 (2.39) | 10.81 (6.13) | 9.33 (4.59) | -1.48 (3.22) | 0.943 | 0.06 |
| **Hostility** | 9.67 (4.46) | 8.19 (4.33) | -1.48 (2.62) | 9.76 (5.25) | 8.29 (4.43) | -1.48 (3.25) | 0.462 | 0.13 |
| **Guilt** | 15.52 (6.49) | 11.1 (5.87) | -4.43 (3.71) | 13.86 (6.80) | 10.95 (6.32) | -2.90 (3.02) | 0.152 | -0.26 |
| **Sadness** | 12.71 (5.61) | 10.48 (5.32) | -2.24 (1.89) | 11.62 (5.10) | 10.05 (5.27) | -1.57 (1.75) | 0.377 | 0.00 |
| **Joviality** | 12.33 (4.40) | 13.62 (5.07) | 1.29 (3.94) | 15.43 (5.57) | 14.62 (6.1) | -0.81 (3.50) | **0.041** | **0.60** |
| **Self-assurance** | 9.67 (4.08) | 10.05 (3.71) | 0.38 (2.27) | 10.48 (4.33) | 10.62 (4.55) | 0.14 (2.10) | 0.736 | 0.13 |
| **Attentiveness** | 9.19 (2.64) | 9.38 (2.84) | 0.19 (2.23) | 10.1 (3.42) | 10.05 (3.60) | -0.05 (1.66) | 0.720 | 0.06 |
| **Shyness** | 8.14 (4.50) | 7.14 (3.54) | -1.00 (1.82) | 7.19 (3.06) | 6.67 (3.34) | -0.52 (1.60) | 0.396 | -0.18 |
| **Fatigue** | 11.86 (4.53) | 11.05 (3.83) | -0.81 (3.03) | 10.43 (4.04) | 9.48 (3.57) | -0.95 (2.29) | 0.839 | 0.05 |
| **Serenity** | 7.10 (1.97) | 8.29 (2.97) | 1.19 (1.91) | 7.67 (2.92) | 8.38 (3.01) | 0.71 (2.12) | 0.427 | 0.14 |
| **Surprise** | 3.52 (1.12) | 4.43 (1.96) | 0.90 (1.64) | 5.00 (3.24) | 5.05 (2.92) | 0.05 (1.53) | 0.057 | 0.54 |

BSRI: Brief State Rumination Inventory; PANASX: Positive Affect and Negative Affect State-Extended

**Extended Data Table 3.** Correlations with Thalamus-vmPFC Connectivity after the intervention.

| **ΔScores (Post-Pre)** | **LIFU** |  | **Sham** |  |
| --- | --- | --- | --- | --- |
|  | **Pearson's r** | **p-value** | **Pearson's r** | **p-value** |
| BSRI | -0.32 | 0.165 | 0.16 | 0.501 |
| PANASX |  |  |  |  |
| Negative affect | -0.03 | 0.884 | -0.28 | 0.235 |
| Positive affect | 0.10 | 0.682 | 0.29 | 0.213 |
| Fear | 0.15 | 0.538 | -0.27 | 0.249 |
| Hostility | -0.17 | 0.470 | -0.32 | 0.166 |
| Guilt | -0.09 | 0.696 | -0.10 | 0.677 |
| Sadness | 0.04 | 0.873 | -0.05 | 0.830 |
| Joviality | 0.06 | 0.797 | 0.34 | 0.145 |
| Self-assurance | 0.03 | 0.913 | 0.15 | 0.518 |
| Attentiveness | 0.19 | 0.423 | 0.40 | 0.082 |
| Shyness | 0.29 | 0.222 | 0.21 | 0.377 |
| Fatigue | -0.21 | 0.376 | -0.32 | 0.176 |
| Serenity | **0.55** | **0.013** | 0.10 | 0.680 |
| Surprise | -0.13 | 0.580 | -0.10 | 0.673 |

**Correlations with Thalamus-PCC Connectivity after the intervention**
| **ΔScores (Post-Pre)** | **LIFU** |  | **Sham** |  |
| --- | --- | --- | --- | --- |
|  | **Pearson's r** | **p-value** | **Pearson's r** | **p-value** |
| BSRI | -0.05 | 0.820 | -0.49 | 0.029 |
| PANASX |  |  |  |  |
| Negative affect | 0.32 | 0.171 | -0.30 | 0.200 |
| Positive affect | -0.19 | 0.412 | 0.41 | 0.073 |
| Fear | 0.19 | 0.430 | -0.27 | 0.254 |
| Hostility | 0.25 | 0.290 | -0.26 | 0.259 |
| Guilt | 0.11 | 0.646 | -0.32 | 0.170 |
| Sadness | 0.13 | 0.599 | -0.26 | 0.260 |
| Joviality | -0.11 | 0.640 | 0.30 | 0.191 |
| Self-assurance | -0.11 | 0.642 | 0.13 | 0.588 |
| Attentiveness | -0.07 | 0.776 | **0.59** | **0.006** |
| Shyness | 0.33 | 0.150 | -0.06 | 0.796 |
| Fatigue | -0.21 | 0.372 | -0.13 | 0.597 |
| Serenity | 0.33 | 0.154 | **0.51** | **0.022** |
| Surprise | 0.31 | 0.184 | 0.12 | 0.608 |

BSRI: Brief State Rumination Inventory; PANASX: Positive Affect and Negative Affect State-Extended

**Extended Data Table 4.** Correlations with Thalamus-vmPFC Connectivity after the intervention.

| **ΔHRV (Post-Pre)** | **LIFU** |  | **Sham** |  |
| --- | --- | --- | --- | --- |
|  | **Pearson's r** | **p-value** | **Pearson's r** | **p-value** |
| SDNN | -0.17 | 0.462 | -0.08 | 0.746 |
| LF | -0.06 | 0.797 | 0.20 | 0.394 |
| HF | 0.03 | 0.889 | -0.29 | 0.212 |
| LF/HF | -0.20 | 0.405 | **0.46** | **0.039** |

**Correlations with Thalamus-PCC Connectivity after the intervention**
| **ΔHRV (Post-Pre)** | **LIFU** |  | **Sham** |  |
| --- | --- | --- | --- | --- |
|  | **Pearson's r** | **p-value** | **Pearson's r** | **p-value** |
| SDNN | **-0.46** | **0.040** | -0.15 | 0.516 |
| LF | 0.17 | 0.482 | 0.38 | 0.099 |
| HF | 0.41 | 0.072 | -0.20 | 0.388 |
| LF/HF | -0.17 | 0.465 | 0.30 | 0.201 |

SDNN: Standard Deviation of Normal Heartbeat Intervals; LF: Low Frequency, HF: High Frequency; LF/HF: Ratio of Low Frequency over High Frequency

**Extended Data Figure 1.**
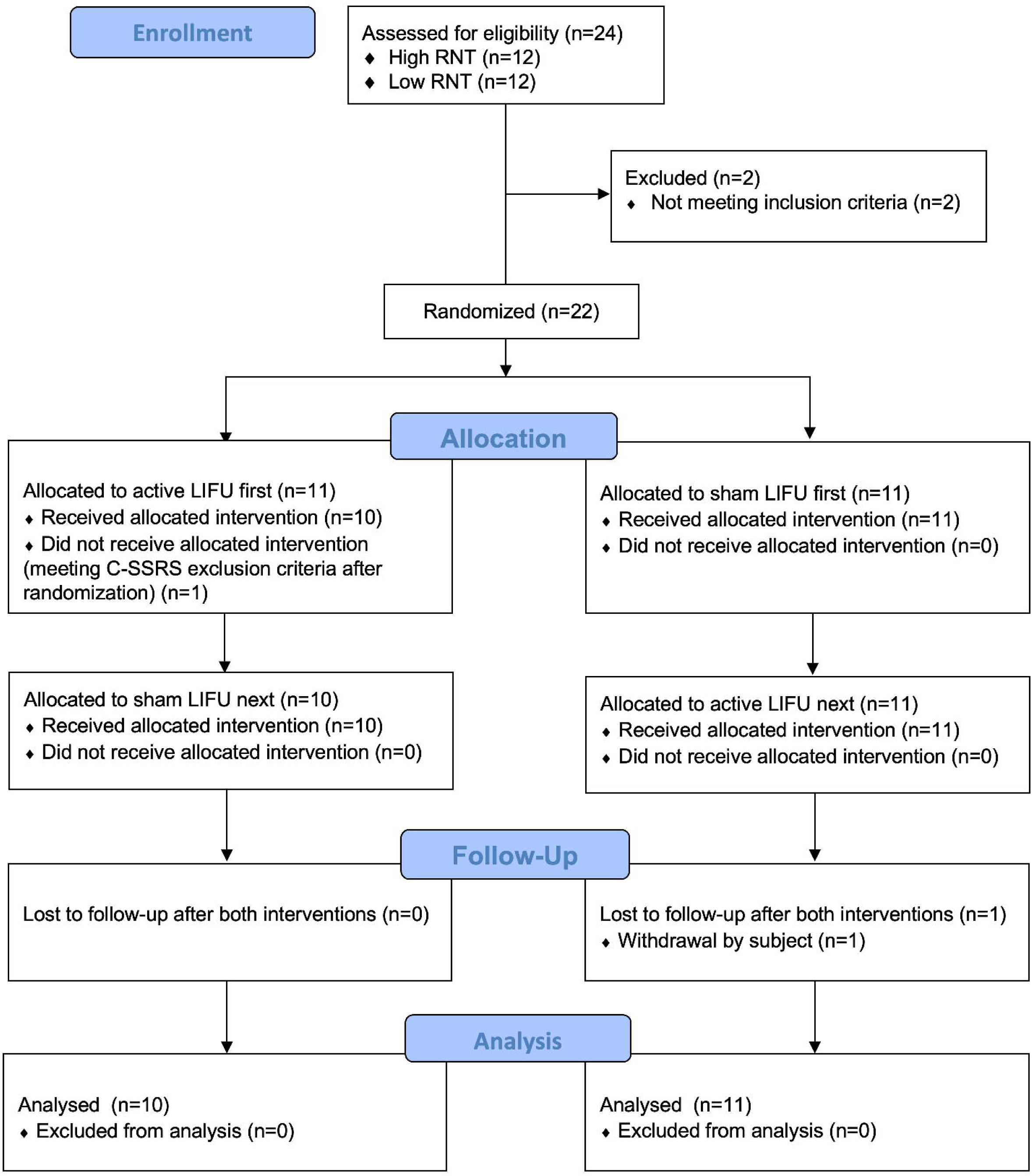
CONSORT diagram of participant flow through the study. A total of 24 participants were assessed for eligibility, with 12 having high repetitive negative thinking (RNT) and 12 having low RNT. Two participants were excluded for not meeting inclusion criteria, and 22 participants were randomized. Each group (n=11) was allocated to either active LIFU first or sham LIFU first. One participant did not receive the active LIFU intervention due to meeting C-SSRS exclusion criteria after randomization. After cross-over, 10 participants from the active-first group and 11 from the sham-first group received both interventions. One participant from the sham-first group withdrew before follow-up. All participants who received the interventions were included in the final analysis.

**Extended Data Figure 2.**
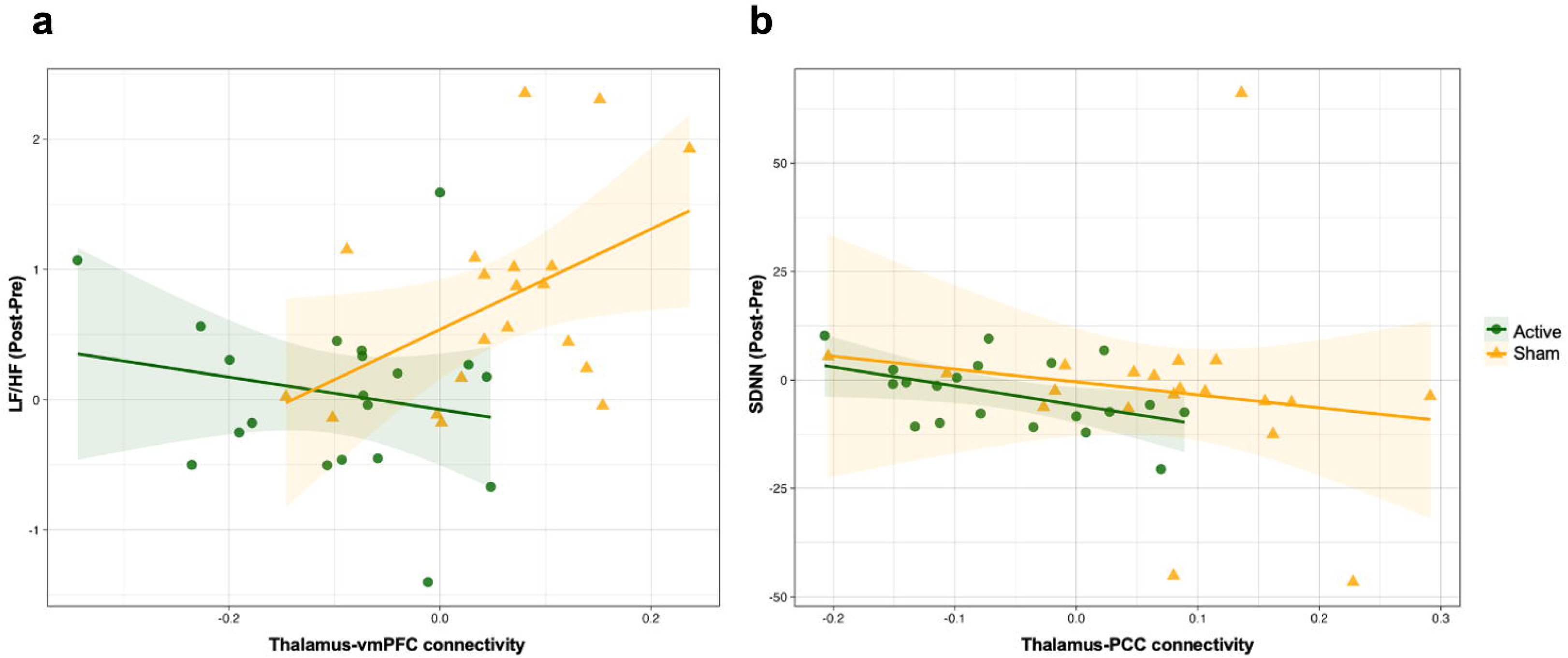
Correlations between changes in resting-state functional connectivity and changes in heart rate variability (HRV). Increased thalamus-vmPFC connectivity is associated with higher sympatho-vagal balance (LF/HF ratio) in the sham group (orange line) but not in the active LIFU group (green line) (a). Reduced thalamus-PCC connectivity correlates with increased overall heart rate variability (SDNN) in the active LIFU group, while this relationship is not observed in the sham group (b).

