## Supplemental materials for "Reversible and Noninvasive Modulation of a Historical Surgical Target for Depression with Low Intensity Focused Ultrasound"

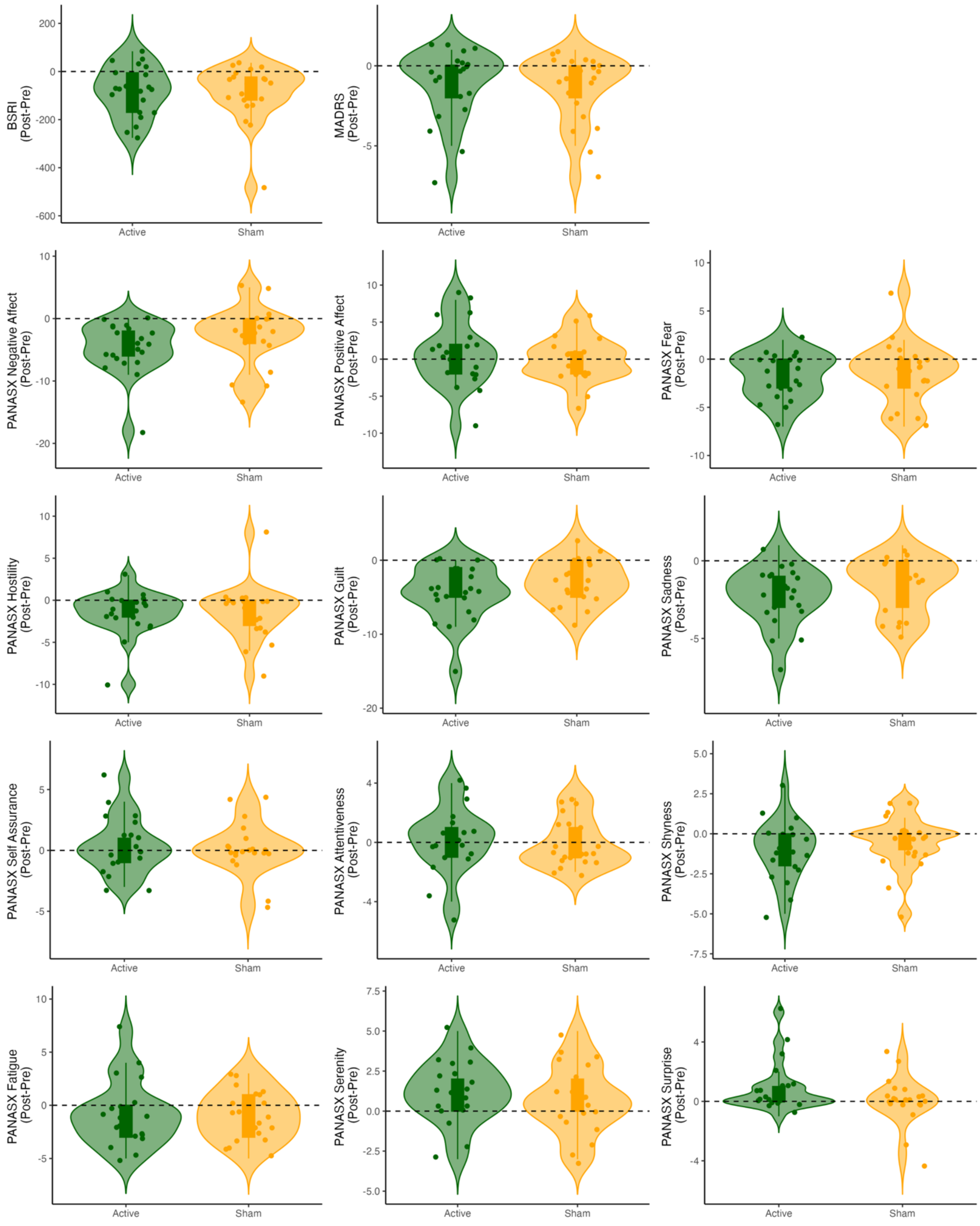


**Supplementary Figure S1. Changes in symptom and mood scales.** Violin plots describing individual and group-level changes in symptom and mood scales for both active LIFU and sham conditions. Each plot compares the active (green) and sham (yellow) conditions, with dots representing individual participant scores, dashed lines indicating median values, and the overall shape representing the distribution of data. BSRI: Brief State Rumination Inventory; MADRS: Montgomery-Asberg Depression Rating Scale; PANASX: Positive and Negative Affective Schedule-Extended.

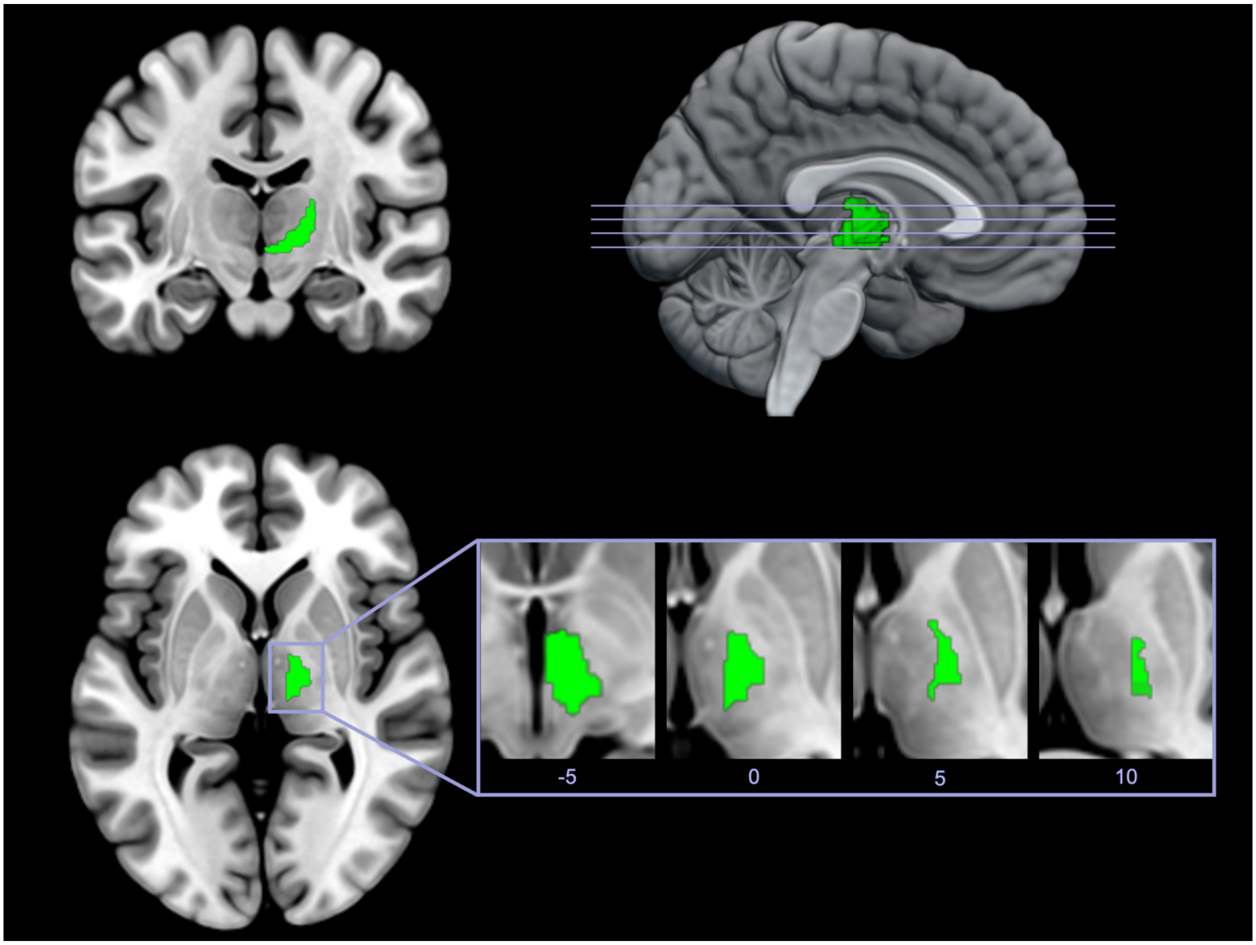


**Supplementary Figure S2**. **Right anterior-ventral thalamus seed region for connectivity analysis.** The right anterior-ventral thalamus region was used as a seed for the seed-to-whole brain connectivity analysis. The seed was defined using the Brainnetome atlas, focusing on thalamic nuclei known for reciprocal connections with the prefrontal cortex, including anterior, ventrolateral, ventromedial, ventral posterior-medial, and ventral posterior-lateral nuclei. This region was used to investigate connectivity changes between active and sham LIFU conditions.
